# Telemedicine Use Among People with HIV in 2021: The Hybrid-Care Environment

**DOI:** 10.1101/2022.06.27.22276960

**Authors:** Walid G. El-Nahal, Geetanjali Chander, Joyce L. Jones, Anthony T. Fojo, Jeanne C. Keruly, Yukari C. Manabe, Richard D. Moore, Kelly A. Gebo, Catherine R. Lesko

## Abstract

**Background:** Telemedicine use for the care of people with HIV (PWH) was widely expanded during the COVID-19 pandemic. During 2021, as on-site care was re-introduced, care was delivered through a mixture of in-person and telemedicine. We studied how different patient populations used telemedicine in this hybrid-care environment.

**Methods:** Using observational data from patients enrolled in the Johns Hopkins HIV Clinical Cohort, we analyzed all in-person and telemedicine HIV primary care visits completed in an HIV clinic from January 1^st^, 2021 to December 30^th^, 2021. We used log-binomial regression models to investigate the association between patient characteristics and the probability of completing a telemedicine versus in-person visit. A secondary analysis of telemedicine visits investigated the probably of completing a video versus telephone visit.

**Results:** A total of 5,518 visits were completed by 1,884 patients; 4,282 (77.6%) visits were in-person, 800 (14.5%) by phone, and 436 (7.9%) by video. The relative risk (RR) of completing telemedicine vs. in-person visits was 0.65 (95% Confidence Interval (CI): 0.47, 0.91) for patients age 65+ vs. age 20-39; 0.84 (95% CI: 0.72, 0.98) for males vs. females; 0.81 (95% CI: 0.66, 0.99) for Black vs. white patients; 0.62 (95% CI: 0.49, 0.79) for patients in the highest vs. lowest quartile of Area Deprivation Index; and 1.52 (95% CI: 1.26, 1.84) for patients >15 miles vs. <5 miles from clinic.

**Conclusions:** In the second year of the pandemic, overall in-person care was utilized more than telemedicine, and significant differences persist across subgroups in telemedicine uptake.

## Introduction

Consistent long-term engagement is crucial to HIV care because it provides access to antiretroviral therapy which dramatically lowers mortality^1–9^ and transmission.^10–13^ At the onset of the COVID-19 pandemic, telemedicine emerged as the primary means of engaging people with HIV (PWH), to mitigate SARS-COV-2 exposure.^14–18^ Later in the pandemic, most clinics resumed more onsite visits and offered a mixture of in-person and telemedicine care,^15^ creating an opportunity to study telemedicine’s potential as a tool for long-term engagement in a hybrid care environment.

Telemedicine’s effect on engagement and treatment has been of great interest during the pandemic.^19– 22^ Early in the pandemic, patients were more likely to complete visits using telemedicine than they had been in-person prior to the pandemic.^14,16^ However, telemedicine uptake varied across demographic subgroups and a significant portion of patients were limited to using telephone-only encounters.^14,15,17,18,23^ Additional data to determine whether these early pandemic trends persisted are needed.

In this study, we investigate how telemedicine use evolved during the second year of the pandemic, characterizing how different patient groups engaged in care when both in-person and telemedicine were widely available. We describe the distribution of (1) in-person versus telemedicine visits and (2) video versus telephone visits utilized by a cohort of people with HIV in care in 2021, and investigate patient characteristics associated with the use of each modality.

## Methods

### Study Sample

The John G. Bartlett Specialty Practice is a large HIV and Hepatitis C subspecialty clinic affiliated with the Johns Hopkins Hospital in East Baltimore. Adults (≥18 years old) with HIV who engage in continuity care at the clinic and consent to share their data are enrolled in the Johns Hopkins HIV Clinical Cohort (JHHCC).^14,24^ Briefly, the JHHCC collects self-reported data including age, gender, race, ethnicity, ZIP code, and HIV acquisition risk factors, as well as electronic medical record data including clinic visits, visit modality (in-person vs. video vs. telephone), lab data, clinical diagnoses, and prescribed treatments. We included all completed HIV primary care visits in the JHHCC from January 1, 2021 through December 31, 2021.

At the onset of the COVID-19 pandemic, starting 3/16/2020, the John G. Bartlett Specialty Practice converted almost all patient encounters to telemedicine. For telemedicine visits, all patients were offered an audio-video encounter initially, with a telephone-only encounter as an alternative if they did not have access to video, declined a video connection, or were unable to connect. All video or telephone visits analyzed herein were full encounters, with all elements of an onsite patient visit except a physical exam. Starting on July 1^st^ 2020, the clinic re-introduced in-person care (aiming for 25% of visits to be in-person initially) and gradually increased it thereafter such that patients were engaging in a mixture of in-person and telemedicine visits. By April 19^th^, 2021, all adults nationwide were eligible for COVID-19 vaccination,^25^ facilitating broader availability of in-person care. By this point, the clinic was encouraging in-person visits, but telemedicine visits were still available for patients who requested them. There was a brief return to telemedicine-preferred visits in late December 2021 coinciding with the COVID-19 Omicron variant surge.

### Outcome of Interest

Our primary outcome was the modality by which a visit was completed: in person or telemedicine. A secondary analysis restricted to telemedicine visits also used a binary outcome: video or telephone.

### Exposures of Interest

We investigated patient characteristics that may be associated with visit modality: age, race, ethnicity, HIV risk factor, recent use of heroin or cocaine use, recent hazardous alcohol use, viral suppression status prior to the study period, history of missed in-person visits, proximity to the clinic, Area Deprivation Index of a patient’s primary residence at the time of the visit, and type of insurance/payor billed for each visit. Age groups were defined as 20-39, 40-64, or ≥65 years-old on January 1^st^, 2021. We combined self-reported race and ethnicity into categories: white, Black, Hispanic, or other/unknown. We categorized HIV risk factors as: patients with high-risk heterosexual intercourse and no other risk factors, men who have sex with men (MSM) with no history of injection drug use (IDU), and patients with a history of IDU regardless of other risk factors. Recent heroin, cocaine, or hazardous alcohol use were determined by trained chart abstractors evaluating medical record notes, labs, and diagnosis codes at 6-month intervals. We defined them using two binary variables: cocaine or heroin use identified in the 6 months before the visit or none; and hazardous alcohol use in the 6 months before the visit or none. We defined prior viral suppression as a viral load <200 copies/mL on the most recent check in the year prior to January 1^st^, 2021. Those with >200 copies/mL on most recent check or no viral loads in 2020 were labeled not suppressed. We defined missed visits as visits that were scheduled in 2021 but neither completed nor cancelled i.e., a no-show visit. Patients were categorized as having no missed visits, 1-2 missed visits or ≥3 missed visits in 2021. Proximity to clinic was calculated using CDX Technologies ZIP Code Distance Batch Report.^26^ It was measured as the distance between the centroid each patient’s 5-digit ZIP code and the centroid of the clinic ZIP code, then categorized into <5 miles, 5-10 miles, 10-15 miles, and ≥15 miles. Insurance status at the time of the visit was categorized into (1) Medicaid or Ryan White, (2) Medicare, (3) private insurance, or (4) other/unknown. The Area Deprivation Index (ADI) was used as another surrogate for socioeconomic status.^27–30^ This index uses information about the poverty rates, educational attainment, employment levels, and housing quality to assign a deprivation ranking to neighborhoods, with higher ADI corresponding to more deprivation. The ADI of each ZIP code was estimated using the median of ADIs of all 9-digit ZIP codes within a patient’s 5-digit ZIP code. We grouped patient ADI values (which range from 1-100) into quartiles.

### Statistical Analysis

We describe the number and proportion of patients that completed each visit type, by patient characteristic. We then used bivariable log-binomial regression models, with one dependent and one independent variable, to investigate the association between each patient characteristic described above and the probability of completing a telemedicine visit versus an in-person visit. In the secondary analysis we restricted to only telemedicine visits and analyzed the probability of completing a video visit versus a telephone visit by patient characteristic.

Our unit of analysis was individual visits. To account for potential correlated outcomes across patients or across providers, we estimated the models using multiway, non-nested clustering by patient and provider with robust standard errors.^31^ After our bivariable analyses, we identified several exposures that appeared strongly correlated with telemedicine and/or video use, and conducted an adjusted supplementary analysis that included terms for race/ethnicity, ADI, and distance from clinic.

## Results

The study population included 1,884 patients who completed a total of 5,518 HIV primary care visits. Of all HIV primary care visits completed, 4,282 (77.6%) were in-person, 800 (14.5%) were by phone only, and 436 (7.9%) were by video. The majority of visits were completed by patients who were age 40-64 (64.2%), male (63.1%), and Black (77.1%); and a plurality identified high-risk heterosexual intercourse as an HIV risk factor (41.6%) (Table 1).

**Table 1.**
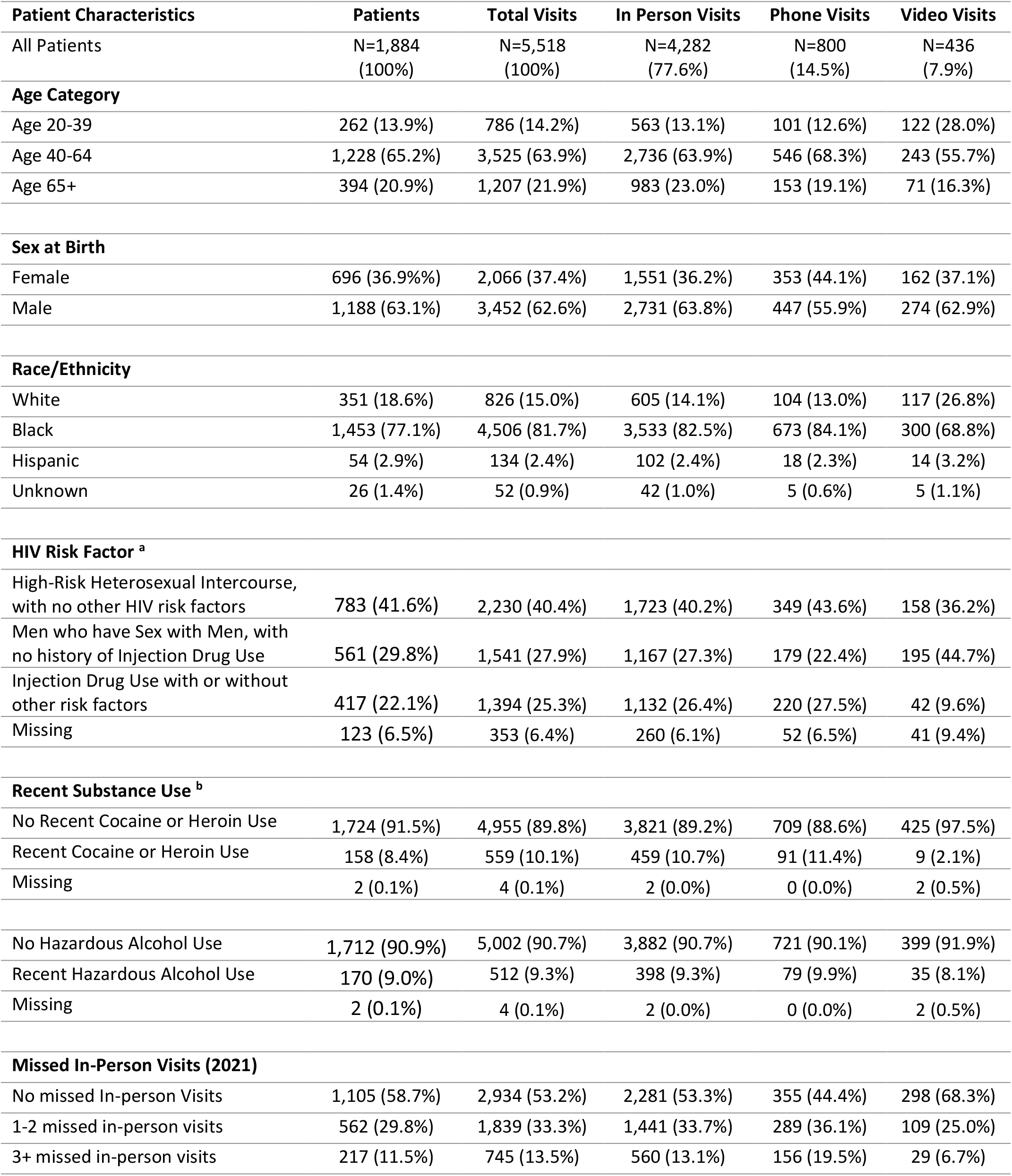
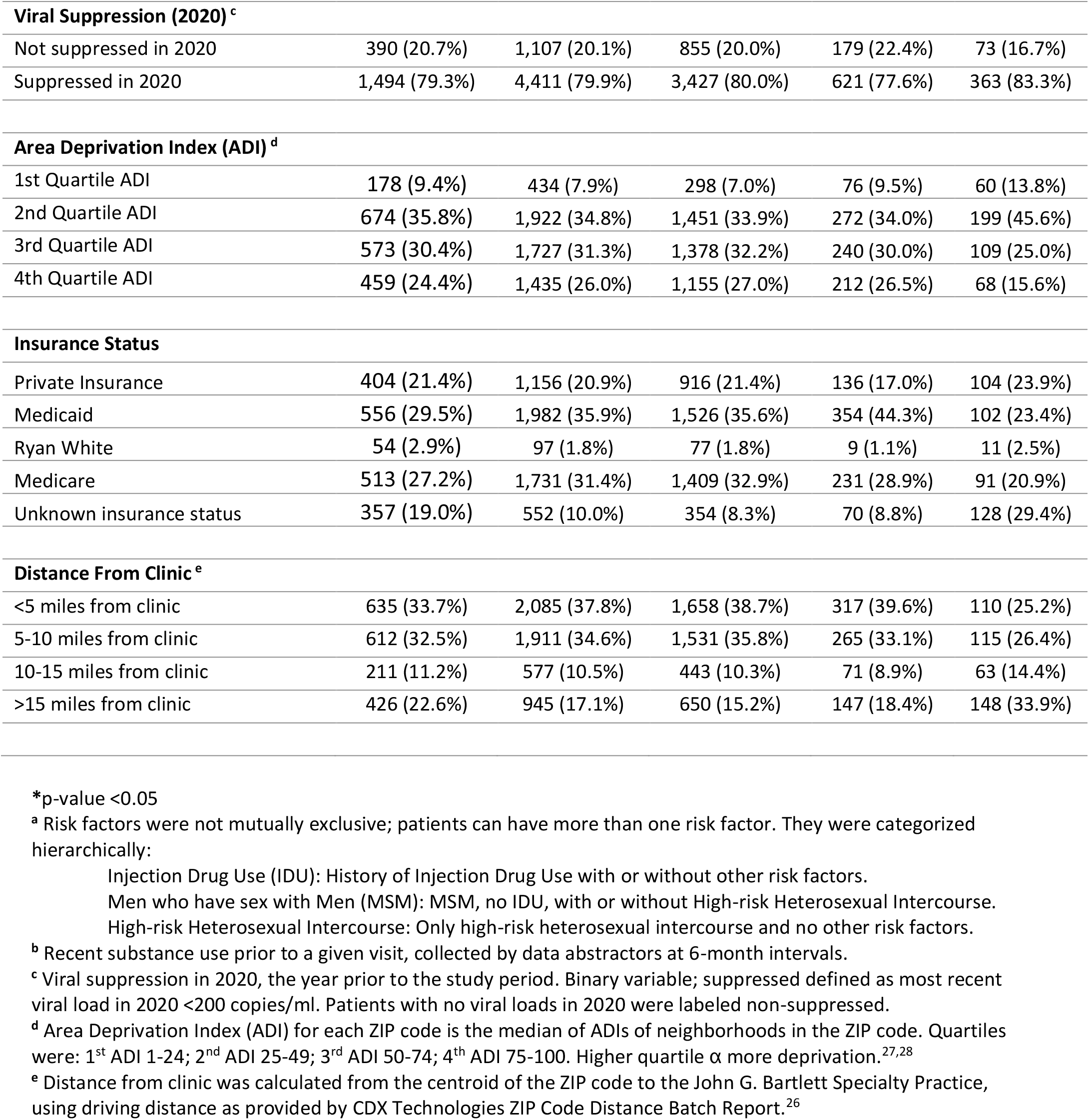
Total Number of Patients, Visits and Visits of Each Type by Patient Characteristics during.

| Patient Characteristics | Patients | Total Visits | In Person Visits | Phone Visits | Video Visits |
| --- | --- | --- | --- | --- | --- |
| All Patients | N=1,884<br>(100%) | N=5,518<br>(100%) | N=4,282<br>(77.6%) | N=800<br>(14.5%) | N=436<br>(7.9%) |
| **Age Category** |  |  |  |  |  |
| Age 20-39 | 262 (13.9%) | 786 (14.2%) | 563 (13.1%) | 101 (12.6%) | 122 (28.0%) |
| Age 40-64 | 1,228 (65.2%) | 3,525 (63.9%) | 2,736 (63.9%) | 546 (68.3%) | 243 (55.7%) |
| Age 65+ | 394 (20.9%) | 1,207 (21.9%) | 983 (23.0%) | 153 (19.1%) | 71 (16.3%) |
| **Sex at Birth** |  |  |  |  |  |
| Female | 696 (36.9%) | 2,066 (37.4%) | 1,551 (36.2%) | 353 (44.1%) | 162 (37.1%) |
| Male | 1,188 (63.1%) | 3,452 (62.6%) | 2,731 (63.8%) | 447 (55.9%) | 274 (62.9%) |
| **Race/Ethnicity** |  |  |  |  |  |
| White | 351 (18.6%) | 826 (15.0%) | 605 (14.1%) | 104 (13.0%) | 117 (26.8%) |
| Black | 1,453 (77.1%) | 4,506 (81.7%) | 3,533 (82.5%) | 673 (84.1%) | 300 (68.8%) |
| Hispanic | 54 (2.9%) | 134 (2.4%) | 102 (2.4%) | 18 (2.3%) | 14 (3.2%) |
| Unknown | 26 (1.4%) | 52 (0.9%) | 42 (1.0%) | 5 (0.6%) | 5 (1.1%) |
| **HIV Risk Factor <sup>a</sup>** |  |  |  |  |  |
| High-Risk Heterosexual Intercourse, with no other HIV risk factors | 783 (41.6%) | 2,230 (40.4%) | 1,723 (40.2%) | 349 (43.6%) | 158 (36.2%) |
| Men who have Sex with Men, with no history of Injection Drug Use | 561 (29.8%) | 1,541 (27.9%) | 1,167 (27.3%) | 179 (22.4%) | 195 (44.7%) |
| Injection Drug Use with or without other risk factors | 417 (22.1%) | 1,394 (25.3%) | 1,132 (26.4%) | 220 (27.5%) | 42 (9.6%) |
| Missing | 123 (6.5%) | 353 (6.4%) | 260 (6.1%) | 52 (6.5%) | 41 (9.4%) |
| **Recent Substance Use <sup>b</sup>** |  |  |  |  |  |
| No Recent Cocaine or Heroin Use | 1,724 (91.5%) | 4,955 (89.8%) | 3,821 (89.2%) | 709 (88.6%) | 425 (97.5%) |
| Recent Cocaine or Heroin Use | 158 (8.4%) | 559 (10.1%) | 459 (10.7%) | 91 (11.4%) | 9 (2.1%) |
| Missing | 2 (0.1%) | 4 (0.1%) | 2 (0.0%) | 0 (0.0%) | 2 (0.5%) |
| No Hazardous Alcohol Use | 1,712 (90.9%) | 5,002 (90.7%) | 3,882 (90.7%) | 721 (90.1%) | 399 (91.9%) |
| Recent Hazardous Alcohol Use | 170 (9.0%) | 512 (9.3%) | 398 (9.3%) | 79 (9.9%) | 35 (8.1%) |
| Missing | 2 (0.1%) | 4 (0.1%) | 2 (0.0%) | 0 (0.0%) | 2 (0.5%) |
| **Missed In-Person Visits (2021)** |  |  |  |  |  |
| No missed In-person Visits | 1,105 (58.7%) | 2,934 (53.2%) | 2,281 (53.3%) | 355 (44.4%) | 298 (68.3%) |
| 1-2 missed in-person visits | 562 (29.8%) | 1,839 (33.3%) | 1,441 (33.7%) | 289 (36.1%) | 109 (25.0%) |
| 3+ missed in-person visits | 217 (11.5%) | 745 (13.5%) | 560 (13.1%) | 156 (19.5%) | 29 (6.7%) |
| **Viral Suppression (2020) <sup>c</sup>** |  |  |  |  |  |
| Not suppressed in 2020 | 390 (20.7%) | 1,107 (20.1%) | 855 (20.0%) | 179 (22.4%) | 73 (16.7%) |
| Suppressed in 2020 | 1,494 (79.3%) | 4,411 (79.9%) | 3,427 (80.0%) | 621 (77.6%) | 363 (83.3%) |
| **Area Deprivation Index (ADI) <sup>d</sup>** |  |  |  |  |  |
| 1st Quartile ADI | 178 (9.4%) | 434 (7.9%) | 298 (7.0%) | 76 (9.5%) | 60 (13.8%) |
| 2nd Quartile ADI | 674 (35.8%) | 1,922 (34.8%) | 1,451 (33.9%) | 272 (34.0%) | 199 (45.6%) |
| 3rd Quartile ADI | 573 (30.4%) | 1,727 (31.3%) | 1,378 (32.2%) | 240 (30.0%) | 109 (25.0%) |
| 4th Quartile ADI | 459 (24.4%) | 1,435 (26.0%) | 1,155 (27.0%) | 212 (26.5%) | 68 (15.6%) |
| **Insurance Status** |  |  |  |  |  |
| Private Insurance | 404 (21.4%) | 1,156 (20.9%) | 916 (21.4%) | 136 (17.0%) | 104 (23.9%) |
| Medicaid | 556 (29.5%) | 1,982 (35.9%) | 1,526 (35.6%) | 354 (44.3%) | 102 (23.4%) |
| Ryan White | 54 (2.9%) | 97 (1.8%) | 77 (1.8%) | 9 (1.1%) | 11 (2.5%) |
| Medicare | 513 (27.2%) | 1,731 (31.4%) | 1,409 (32.9%) | 231 (28.9%) | 91 (20.9%) |
| Unknown insurance status | 357 (19.0%) | 552 (10.0%) | 354 (8.3%) | 70 (8.8%) | 128 (29.4%) |
| **Distance From Clinic <sup>e</sup>** |  |  |  |  |  |
| <5 miles from clinic | 635 (33.7%) | 2,085 (37.8%) | 1,658 (38.7%) | 317 (39.6%) | 110 (25.2%) |
| 5-10 miles from clinic | 612 (32.5%) | 1,911 (34.6%) | 1,531 (35.8%) | 265 (33.1%) | 115 (26.4%) |
| 10-15 miles from clinic | 211 (11.2%) | 577 (10.5%) | 443 (10.3%) | 71 (8.9%) | 63 (14.4%) |
| >15 miles from clinic | 426 (22.6%) | 945 (17.1%) | 650 (15.2%) | 147 (18.4%) | 148 (33.9%) |

\*p-value <0.05
<sup>a</sup> Risk factors were not mutually exclusive; patients can have more than one risk factor. They were categorized hierarchically:
Injection Drug Use (IDU): History of Injection Drug Use with or without other risk factors.
Men who have sex with Men (MSM): MSM, no IDU, with or without High-risk Heterosexual Intercourse.
High-risk Heterosexual Intercourse: Only high-risk heterosexual intercourse and no other risk factors.
<sup>b</sup> Recent substance use prior to a given visit, collected by data abstractors at 6-month intervals.
<sup>c</sup> Viral suppression in 2020, the year prior to the study period. Binary variable; suppressed defined as most recent viral load in 2020 <200 copies/ml. Patients with no viral loads in 2020 were labeled non-suppressed.
<sup>d</sup> Area Deprivation Index (ADI) for each ZIP code is the median of ADIs of neighborhoods in the ZIP code. Quartiles were: 1<sup>st</sup> ADI 1-24; 2<sup>nd</sup> ADI 25-49; 3<sup>rd</sup> ADI 50-74; 4<sup>th</sup> ADI 75-100. Higher quartile α more deprivation. <sup>27,28</sup>
<sup>e</sup> Distance from clinic was calculated from the centroid of the ZIP code to the John G. Bartlett Specialty Practice, using driving distance as provided by CDX Technologies ZIP Code Distance Batch Report. <sup>26</sup>

The probability of completing a telemedicine visit rather than an in-person visit (Table 2) was 35% lower for patients who were age 65+ compared to Age 20-39 (Risk Ratio (RR): 0.65, 95% Confidence Interval (CI): 0.47, 0.91);16% lower for males compared to females (RR: 0.84, 95% CI: 0.72, 0.98); 19% lower for Black patients compared to white patients (RR: 0.81, 95% CI: 0.66, 0.99); 17% lower for patients with a history of injection drug use compared to those with high-risk heterosexual intercourse (RR: 0.83, 95% CI: 0.71, 0.96); and 38% lower for patients living in a ZIP code in the highest deprivation quartile compared to the lowest deprivation quartile (RR: 0.62, 95% CI: 0.49, 0.79). The probability of completing a telemedicine visit was 52% higher for patients who lived ≥15 miles from the clinic compared to those who lived <5 miles from the clinic (RR: 1.52, 95% CI: 1.26, 1.84). Missed visits, prior viral suppression, insurance status and recent hazardous alcohol use were not significantly associated with differences in telemedicine use.

**Table 2.** Relative Use of Telemedicine vs. In-Person Visits and Video vs. Phone Visits by Patient Characteristic.

| Patient Characteristics | Telemedicine Vs. In-Person |  |  |  | Video vs. Phone |  |  |  |
| --- | --- | --- | --- | --- | --- | --- | --- | --- |
|  | Bivariable Analysis<sup>a</sup> |  | Adjusted Analysis<sup>b</sup> |  | Bivariable Analysis<sup>a</sup> |  | Adjusted Analysis<sup>b</sup> |  |
| Age Category |  |  |  |  |  |  |  |  |
| Age 20-39 | 1.00 | Ref. |  |  | 1.00 | Ref. |  |  |
| Age 40-64 | 0.79 | (0.58-1.08) |  |  | 0.56* | (0.43-0.74) |  |  |
| Age 65+ | 0.65* | (0.47-0.91) |  |  | 0.58* | (0.41-0.83) |  |  |
| Sex at Birth |  |  |  |  |  |  |  |  |
| Female | 1.00 | Ref. |  |  | 1.00 | Ref. |  |  |
| Male | 0.84* | (0.72-0.98) |  |  | 1.21 | (0.93-1.56) |  |  |
| Race/Ethnicity |  |  |  |  |  |  |  |  |
| White | 1.00 | Ref. | 1.00 | Ref. | 1.00 | Ref. | 1.00 | Ref. |
| Black | 0.81* | (0.66-0.99) | 0.95 | (0.78-1.16) | 0.58* | (0.45-0.76) | 0.72* | (0.55-0.96) |
| Hispanic | 0.89 | (0.62-1.29) | 0.98 | (0.68-1.40) | 0.83 | (0.46-1.49) | 0.82 | (0.48-1.40) |
| Race Unknown | 0.72 | (0.39-1.32) | 0.74 | (0.42-1.32) | 0.94 | (0.47-1.89) | 0.85 | (0.43-1.70) |
| HIV Risk Factors <sup>c</sup> |  |  |  |  |  |  |  |  |
| High-Risk Heterosexual Intercourse, with no other HIV risk factors | 1.00 | Ref. |  |  | 1.00 | Ref. |  |  |
| Men who have Sex with Men, with no history of Injection Drug Use | 1.07 | (0.88-1.30) |  |  | 1.67* | (1.35-2.07) |  |  |
| Injection Drug Use with or without other risk factors | 0.83* | (0.71-0.96) |  |  | 0.51* | (0.33-0.80) |  |  |
| Recent Substance Use (6 months) <sup>d</sup> |  |  |  |  |  |  |  |  |
| No Recent Cocaine or Heroin Use | 1.00 | Ref. |  |  | 1.00 | Ref. |  |  |
| Recent Cocaine or Heroin Use | 0.78 | (0.59-1.04) |  |  | 0.24* | (0.13-0.45) |  |  |
| No Recent Hazardous Alcohol Use | 1.00 | Ref. |  |  | 1.00 | Ref. |  |  |
| Recent Hazardous Alcohol use | 0.99 | (0.82-1.21) |  |  | 0.86 | (0.62-1.21) |  |  |
| Missed In-Person Visits in 2021 |  |  |  |  |  |  |  |  |
| No Missed In-Person Visits in 2021 | 1.00 | Ref. |  |  | 1.00 | Ref. |  |  |
| 1-2 Missed In-Person Visits in 2021 | 0.97 | (0.87-1.09) |  |  | 0.60* | (0.45-0.80) |  |  |
| 3+ Missed In-Person Visits in 2021 | 1.12 | (0.87-1.44) |  |  | 0.34* | (0.21-0.56) |  |  |
| Viral Suppression Status in 2020 <sup>e</sup> |  |  |  |  |  |  |  |  |
| Not Suppressed | 1.00 | Ref. |  |  | 1.00 | Ref. |  |  |
| Suppressed | 0.98 | (0.84-1.14) |  |  | 1.27 | (0.97-1.67) |  |  |
| Area Deprivation Index <sup>f</sup> |  |  |  |  |  |  |  |  |
| 1st Quartile ADI | 1.00 | Ref. | 1.00 | Ref. | 1.00 | Ref. | 1.00 | Ref. |
| 2nd Quartile ADI | 0.78* | (0.62-0.99) | 0.88 | (0.70-1.09) | 0.96 | (0.73-1.25) | 1.07 | (0.83-1.38) |
| 3rd Quartile ADI | 0.64* | (0.50-0.83) | 0.81 | (0.63-1.03) | 0.71* | (0.54-0.92) | 0.86 | (0.63-1.18) |
| 4th Quartile ADI | 0.62* | (0.49-0.79) | 0.75* | (0.57-0.98) | 0.55* | (0.39-0.78) | 0.77 | (0.52- 1.12) |

| **Insurance Status <sup>g</sup>** |  |  |  |  |  |  |  |  |
| --- | --- | --- | --- | --- | --- | --- | --- | --- |
| Private Insurance | 1.00 | Ref. |  |  | 1.00 | Ref. |  |  |
| Medicaid or Ryan White | 1.10 | (0.91-1.34) |  |  | 0.55* | (0.45-0.67) |  |  |
| Medicare | 0.90 | (0.71-1.14) |  |  | 0.65* | (0.48-0.89) |  |  |
| **Distance From Clinic <sup>h</sup>** |  |  |  |  |  |  |  |  |
| Less than 5 miles | 1.00 | Ref. | 1.00 | Ref. | 1.00 | Ref. | 1.00 | Ref. |
| Between 5-10 miles | 0.97 | (0.86-1.09) | 0.95 | (0.85-1.08) | 1.17 | (0.90-1.53) | 1.15 | (0.84-1.57) |
| Between 10-15 miles | 1.13 | (0.93-1.38) | 1.06 | (0.87-1.29) | 1.83* | (1.39-2.40) | 1.52* | (1.14-2.02) |
| Greater than 15 miles | 1.52* | (1.26-1.84) | 1.37* | (1.14-1.64) | 1.95* | (1.52-2.50) | 1.52* | (1.14- 2.02) |

\*p-value <0.05.
<sup>a</sup> Bivariable analysis: Independent outcome (1 patient characteristic), Dependent outcome (visit type).
<sup>b</sup> Adjusted analysis: Race/Ethnicity, Area Deprivation Index Category, Distance from clinic, and Dependent Outcome (visit type).
<sup>c</sup> Risk factors were not mutually exclusive; patients can have more than one risk factor. They were categorized hierarchically:
Injection Drug Use (IDU): History of Injection Drug Use with or without other risk factors.
Men who have sex with Men (MSM): MSM, no IDU, with or without High-risk Heterosexual Intercourse.
High-risk Heterosexual Intercourse: Only high-risk heterosexual intercourse and no other risk factors.
<sup>d</sup> Recent substance use prior to a given visit, collected by data abstractors at 6-month intervals.
<sup>e</sup> Viral suppression in 2020, the year prior to the study period. Binary variable; suppressed defined as most recent viral load in 2020 <200 copies/ml. Patients with no viral loads in 2020 were labeled non-suppressed.
<sup>f</sup> Area Deprivation Index (ADI) for each ZIP code is the median of ADIs of neighborhoods in the ZIP code. Quartiles were: 1<sup>st</sup> ADI 1-24; 2<sup>nd</sup> ADI 25-49; 3<sup>rd</sup> ADI 50-74; 4<sup>th</sup> ADI 75-100. Higher quartile α more deprivation.<sup>27,28</sup>
<sup>g</sup> All patients with Medicaid or Ryan White compared to everyone else, then all patients with private insurance compared to everyone else.
<sup>h</sup> Distance from clinic was calculated from the centroid of the ZIP code to the John G. Bartlett Specialty Practice, using driving distance as provided by CDX Technologies ZIP Code Distance Batch Report.<sup>26</sup>

Per the primary findings, the telehealth visits analyzed were completed by a population that was disproportionately younger (18% of visits <age 40), more female (41.7% of visits), had lower rates of IDU (21.2% of visits), and lived in areas that were further from clinic (23.9% of visits >15 miles) with a lower ADI (11% of visits in lowest quartile of deprivation). Among these telehealth visits, the likelihood of using video versus phone (Table 2) was lower among patients who were older, Black, had a history of injection drug use, had recent cocaine or heroin use, missed 3 or more in-person visits in 2021, lived in an area with high deprivation, or relied on Medicaid or Ryan White for their visit. The likelihood of video use was higher for patients with private insurance, patients who lived further from the clinic, or patients who are MSM. Sex at birth, prior viral suppression, and recent hazardous alcohol use, were not significantly associated with differences in video versus telephone use. The likelihood of telemedicine versus in-person care and video versus phone visits for all patient characteristics are presented in Figure 1.

**Figure 1.**
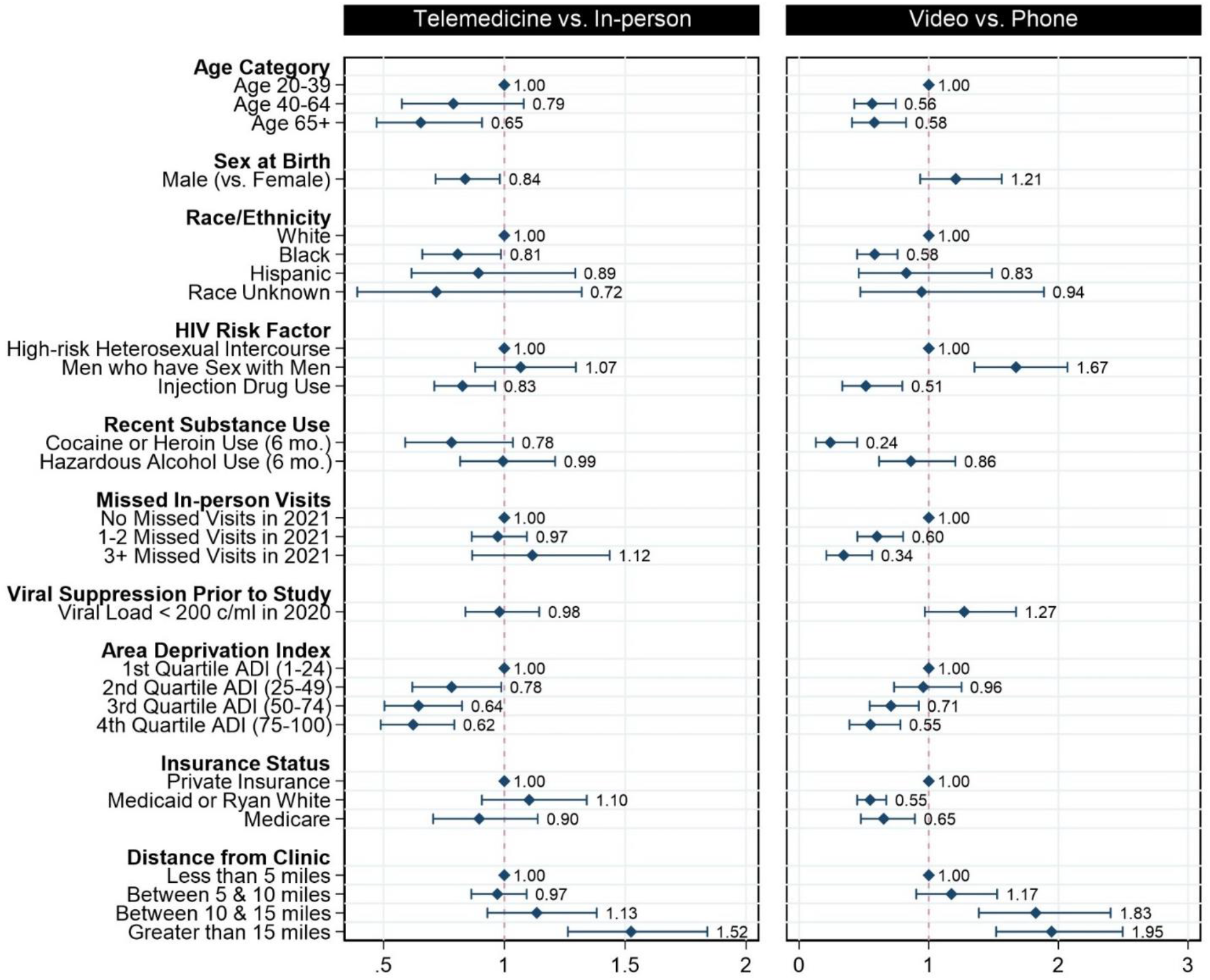
Relative Use of Telemedicine vs. In-Person Visits and Video vs. Phone Visits by Patient Characteristic.

Finally, the adjusted analysis included terms for distance from clinic, area deprivation, race and ethnicity. In this adjusted analysis (Table 2), distance from clinic remained associated with both telemedicine and video use, while higher deprivation was associated with lower likelihood of telemedicine use and Black race with lower likelihood of video use. Each of these associations was attenuated compared to the original analysis.

## Discussion

In this survey of an urban clinic in Baltimore, MD, most HIV primary care visits in 2021 were completed in-person and most visits completed remotely were conducted by telephone rather than video visit.

Patients who are older, male, Black, identify IDU as an HIV risk factor, or live in areas with greater deprivation, were less likely to use telemedicine than others. Patients who are older, Black, identify IDU as an HIV risk factor, have recent cocaine or heroin use, missed multiple in-person visits, are on Medicaid or Ryan White, or live in areas with higher ADIs are less likely to use video compared to telephone when they did complete telemedicine visits. Conversely, patients who have private insurance, live in areas with lower ADIs, live further from clinic or are MSM are more likely to use video for telemedicine visits.

Existing data have characterized telemedicine use for people with HIV early in the pandemic, during a period of peak disruption and blanket telemedicine adoption when remote visits were the main option available.^14,15^ The data herein instead provides insight into telemedicine uptake after in-person visits were widely re-introduced, when patients and providers had more flexibility to choose visit modality. We expect this hybrid-care environment of both remote and in-person visits offers a better representation of how telemedicine may be used long-term beyond the pandemic.

That said, our findings are largely consistent with data from multiple studies conducted earlier in the pandemic.^14,15,17,21,32^ Several groups were less likely to use telemedicine generally and video visits specifically, including older patients and Black patients. While survey data suggest that attitudes towards telemedicine use do not differ across these demographics,^33^ it is hypothesized that this disparity is a consequence of the “digital divide”,^21,32^ structural factors that result in differential computer access, internet access and technological literacy which disproportionately impact older patients and racial minorities.^34–36^ This is consistent with our findings that surrogates for socioeconomic status, including higher deprivation ZIP codes or reliance on Medicaid or Ryan White are also associated with less telemedicine and video use, and that adjusting for ADI attenuated the association of race with likelihood of telemedicine and video use.

Prior suppression status and missed in-person visits were studied to give insight into what visit modalities are used by patients with less consistent engagement and suppression. Patients who miss multiple in-person visits are not more likely to use telemedicine when they do complete a visit, and if they do it is often by telephone. This raises doubts about whether telemedicine does increase access to patients who are less able to consistently engage with care, and warrants further study. Patients with recent cocaine or heroin, another population at risk for continuum-of-care disruptions, were also less likely to use video visits, again consistent with findings from the first months of the pandemic.^14,15^ This remains a critical finding given the ongoing interest in the use of telemedicine for the management of substance use disorders moving forward,^37,38^ as it remains unclear if telephone and video visits offer comparable outcomes.

Several populations were more likely to use telemedicine, specifically video visits. Data from this cohort^14^ and others^15,39–41^ earlier in the pandemic showed that female patients were more likely to use telemedicine than male patients,^14^ a finding that was repeated in this analysis. We previously hypothesized that this may be due to the disproportionate impact of the burden of caregiving and transportation barriers among our female patients.^42,43^ Men who have sex with men were much more likely to use video than patients who identified other HIV risk factors. A higher proportion of MSM in our cohort were white, younger, less likely to use Medicaid, and more likely to live in areas with low deprivation indices, all factors that are associated with video use and may explain this finding. Finally, patients who lived further from the clinic were more likely to use telemedicine, likely due to the challenges of a longer commute. They were also more likely to use video. In a supplementary analysis, that adjusted for several factors associated with video use (distance, ADI, and race/ethnicity), distance from clinic maintained a strong association with video use (Table 1).

There are various implications from these findings. These data reflect telemedicine uptake during a period where patients had a choice between in person and telemedicine visits. In 2021, in-person was more commonly used and differential uptake of telemedicine was observed across subgroups. For those more likely to use telemedicine, it may function as means of increasing access or an alternative for patients with barriers to in-person care. Conversely, there are several groups whose use of telemedicine is lower by comparison, including older patients, Black patients, patients living in high-ADI neighborhoods, patients with Medicaid or Ryan White and patients with substance use disorder. Future studies will need to investigate potential barriers to telemedicine that differentially impact patient access, given that telemedicine use has continued to be a part of care delivery. Use of video vs. telephone for telemedicine is particularly important, because of disparate reimbursement structures that incentivize providing video over telephone visits, restricting access for patients likely to have telephone access only.

Several limitations of this analysis are worth noting. COVID-19 vaccination status may impact a patient’s likelihood of completing an in-person visit, due to concerns about SARS-CoV-2 exposure risk. Vaccine uptake changed significantly during 2021. Less than 1% of the Maryland population was fully vaccinated at the start of 2021, compared to over 70% of state residents by the end of the year,^44^ which may have influenced patient comfort with in-person care over time. Furthermore, these data are limited to a single site at an academic center in an urban setting, which may not be generalizable given that telemedicine implementation varied in HIV clinics across the country. Additionally, while these data speak to how patients engaged in care, it does not give insight into the quality of the care delivered remotely. It remains to be seen if engaging through telemedicine improves downstream outcomes such as viral suppression or comorbidity treatment in the same way in-person care does. Limited early data suggests telemedicine may be comparable to in-person care for achieving viral suppression.^45^ Finally, ADI most accurately maps to a census block group, which is a smaller geographic unit than a ZIP code. Its accuracy for use with ZIP codes has been questioned when validated in the past,^28,46^ however, given the geographic resolution of our data, we used ADI as an imperfect approximation, but this may hinder our ability to fully estimate the associations between deprivation and telemedicine use.

## Conclusions

In the hybrid-care environment where both in-person and remote care are offered, people living with HIV used in-person visits more than telemedicine, but telemedicine continued to be a sizeable portion of visits in the second year of the pandemic. Age, sex, race, surrogates for socioeconomic status, recent substance use, and distance to clinic were all found to be associated with how likely patients were to engage with video and telephone visits. Future work investigating why differential uptake exists across patient populations and whether telemedicine use is associated with comparable outcomes to in-person care is essential. If telemedicine can be made accessible to all, hybrid care may offer a means of increasing access beyond the pandemic and reaching a wider portion of the patient population moving forward.

## Data Availability

All data produced in the present study are available upon reasonable request to the authors.

## Acknowledgements & Funding

All authors have contributed significantly to this work and have approved of the manuscript as submitted. This work was supported by grants from the National Institutes of Health [T32 AI007291, K24 AA027483, K01 AA028193, K08 MH118094, U01 DA036935 and P30 AI094189]. The content is solely the responsibility of the authors and does not necessarily represent the official views of the National Institutes of Health.

## STROBE Statement–checklist of items that should be included in reports of observational studies

STROBE Statement—checklist of items that should be included in reports of observational studies
|  | Item<br>No | Recommendation |
| --- | --- | --- |
| ✓ **Title and abstract** | 1 | (a) Indicate the study's design with a commonly used term in the title or the abstract<br>(b) Provide in the abstract an informative and balanced summary of what was done and what was found |
| ✓ **Introduction** |  |  |
| ✓ Background/rationale | 2 | Explain the scientific background and rationale for the investigation being reported |
| ✓ Objectives | 3 | State specific objectives, including any prespecified hypotheses |
| ✓ **Methods** |  |  |
| ✓ Study design | 4 | Present key elements of study design early in the paper |
| ✓ Setting | 5 | Describe the setting, locations, and relevant dates, including periods of recruitment, exposure, follow-up, and data collection |
| ✓ Participants | 6 | (a) *Cohort study*—Give the eligibility criteria, and the sources and methods of selection of participants. Describe methods of follow-up<br>*Case-control study*—Give the eligibility criteria, and the sources and methods of case ascertainment and control selection. Give the rationale for the choice of cases and controls<br>*Cross-sectional study*—Give the eligibility criteria, and the sources and methods of selection of participants<br>(b) *Cohort study*—For matched studies, give matching criteria and number of exposed and unexposed<br>*Case-control study*—For matched studies, give matching criteria and the number of controls per case |
| ✓ Variables | 7 | Clearly define all outcomes, exposures, predictors, potential confounders, and effect modifiers. Give diagnostic criteria, if applicable |
| ✓ Data sources/<br>measurement | 8* | For each variable of interest, give sources of data and details of methods of assessment (measurement). Describe comparability of assessment methods if there is more than one group |
| ✓ Bias | 9 | Describe any efforts to address potential sources of bias |
| ✓ Study size | 10 | Explain how the study size was arrived at |
| ✓ Quantitative variables | 11 | Explain how quantitative variables were handled in the analyses. If applicable, describe which groupings were chosen and why |
| ✓ Statistical methods | 12 | (a) Describe all statistical methods, including those used to control for confounding |
|  |  | (b) Describe any methods used to examine subgroups and interactions |
|  |  | (c) Explain how missing data were addressed |
|  |  | (d) *Cohort study*—If applicable, explain how loss to follow-up was addressed |
|  |  | *Case-control study*—If applicable, explain how matching of cases and controls was addressed |
|  |  | *Cross-sectional study*—If applicable, describe analytical methods taking account of sampling strategy |
|  |  | (e) Describe any sensitivity analyses |

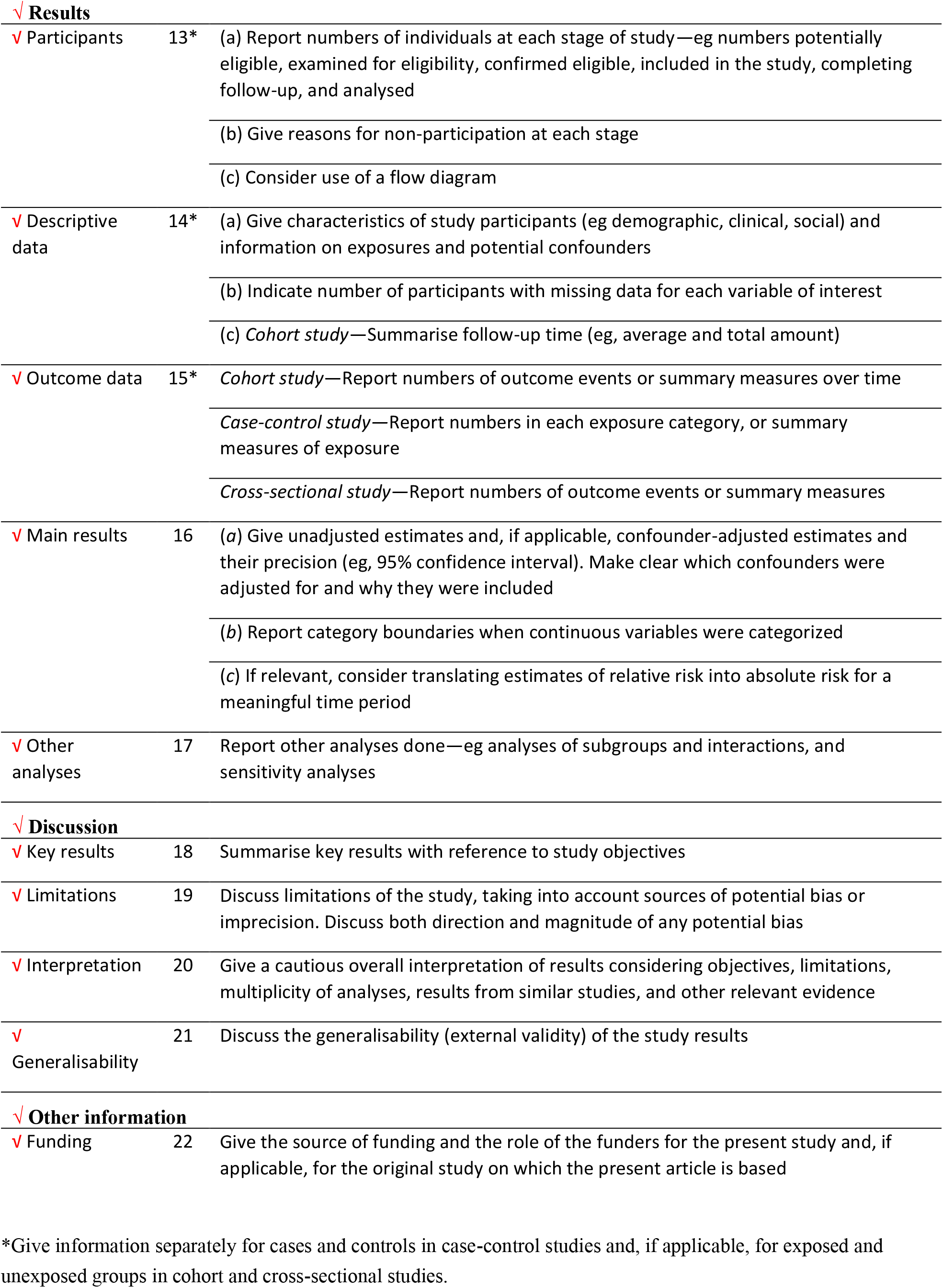
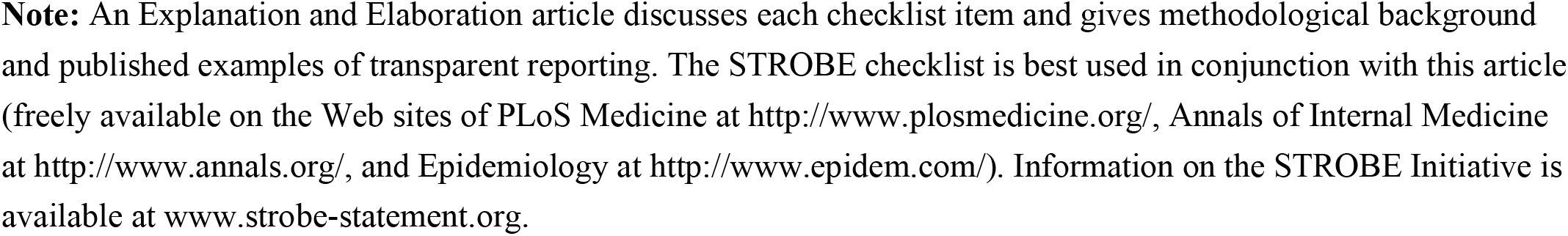

|  |  |  |
| --- | --- | --- |
| **✓ Results** |  |  |
| ✓ Participants | 13* | (a) Report numbers of individuals at each stage of study—eg numbers potentially eligible, examined for eligibility, confirmed eligible, included in the study, completing follow-up, and analysed<br>(b) Give reasons for non-participation at each stage<br>(c) Consider use of a flow diagram |
| ✓ Descriptive data | 14* | (a) Give characteristics of study participants (eg demographic, clinical, social) and information on exposures and potential confounders<br>(b) Indicate number of participants with missing data for each variable of interest<br>(c) *Cohort study*—Summarise follow-up time (eg, average and total amount) |
| ✓ Outcome data | 15* | *Cohort study*—Report numbers of outcome events or summary measures over time<br>*Case-control study*—Report numbers in each exposure category, or summary measures of exposure<br>*Cross-sectional study*—Report numbers of outcome events or summary measures |
| ✓ Main results | 16 | (a) Give unadjusted estimates and, if applicable, confounder-adjusted estimates and their precision (eg, 95% confidence interval). Make clear which confounders were adjusted for and why they were included<br>(b) Report category boundaries when continuous variables were categorized<br>(c) If relevant, consider translating estimates of relative risk into absolute risk for a meaningful time period |
| ✓ Other analyses | 17 | Report other analyses done—eg analyses of subgroups and interactions, and sensitivity analyses |
| **✓ Discussion** |  |  |
| ✓ Key results | 18 | Summarise key results with reference to study objectives |
| ✓ Limitations | 19 | Discuss limitations of the study, taking into account sources of potential bias or imprecision. Discuss both direction and magnitude of any potential bias |
| ✓ Interpretation | 20 | Give a cautious overall interpretation of results considering objectives, limitations, multiplicity of analyses, results from similar studies, and other relevant evidence |
| ✓ Generalisability | 21 | Discuss the generalisability (external validity) of the study results |
| **✓ Other information** |  |  |
| ✓ Funding | 22 | Give the source of funding and the role of the funders for the present study and, if applicable, for the original study on which the present article is based |

\*Give information separately for cases and controls in case-control studies and, if applicable, for exposed and unexposed groups in cohort and cross-sectional studies.
**Note:** An Explanation and Elaboration article discusses each checklist item and gives methodological background and published examples of transparent reporting. The STROBE checklist is best used in conjunction with this article (freely available on the Web sites of PLoS Medicine at http://www.plosmedicine.org/, Annals of Internal Medicine at http://www.annals.org/, and Epidemiology at http://www.epidem.com/). Information on the STROBE Initiative is available at www.strobe-statement.org.

## References

1. Ulloa AC, Puskas C, Yip B, et al. Retention in care and mortality trends among patients receiving comprehensive care for HIV infection: a retrospective cohort study. C Open. 2019;7(2):E236. doi:10.9778/CMAJO.20180136

2. Sabin CA, Howarth A, Jose S, et al. Association between engagement in-care and mortality in HIV-positive persons. AIDS. 2017;31(5):653. doi:10.1097/QAD.0000000000001373

3. Horberg MA, Hurley LB, Silverberg MJ, Klein DB, Quesenberry CP, Mugavero MJ. Missed office visits and risk of mortality among hiv-infected subjects in a large healthcare system in the United States. AIDS Patient Care STDS. 2013;27(8):442–449. doi:10.1089/apc.2013.0073

4. Kay ES, Batey DS, Westfall AO, et al. Compound Retention in Care and All-Cause Mortality Among Persons Living With Human Immunodeficiency Virus. Open Forum Infect Dis. 2019;6(4). doi:10.1093/OFID/OFZ120

5. Mugavero MJ, Westfall AO, Cole SR, et al. Beyond Core Indicators of Retention in HIV Care: Missed Clinic Visits Are Independently Associated With All-Cause Mortality. Clin Infect Dis. 2014;59(10):1471–1479. doi:10.1093/CID/CIU603

6. Edwards JK, Cole SR, Breger TL, et al. Mortality Among Persons Entering HIV Care Compared With the General U.S. Population. https://doi.org/107326/M21-0065. 2021;174(9):p1197-1206. doi:10.7326/M21-0065

7. Poorolajal J, Hooshmand E, Mahjub H, Esmailnasab N, Jenabi E. Survival rate of AIDS disease and mortality in HIV-infected patients: a meta-analysis. Public Health. 2016;139:3–12. doi:10.1016/j.puhe.2016.05.004

8. Mocroft A, Ledergerber B, Katlama C, et al. Decline in the AIDS and death rates in the EuroSIDA study: an observational study. Lancet. 2003;362(9377):22–29. doi:10.1016/S0140-6736(03)13802-0

9. Palella FJ, Delaney KM, Moorman AC, et al. Declining Morbidity and Mortality among Patients with Advanced Human Immunodeficiency Virus Infection. N Engl J Med. 1998;338(13):853–860. doi:10.1056/NEJM199803263381301

10. Cohen MS, Chen YQ, McCauley M, et al. Antiretroviral Therapy for the Prevention of HIV-1 Transmission. N Engl J Med. 2016;375(9):830–839. doi:10.1056/NEJMOA1600693/SUPPL_FILE/NEJMOA1600693_DISCLOSURES.PDF

11. Shah M, Perry A, Risher K, et al. Effect of the US National HIV/AIDS Strategy targets for improved HIV care engagement: A modelling study. Lancet HIV. 2016;3(3):e140–e146. doi:10.1016/S2352-3018(16)00007-2

12. Li Z, Purcell DW, Sansom SL, Hayes D, Hall HI. Vital Signs: HIV Transmission Along the Continuum of Care — United States, 2016. MMWR Morb Mortal Wkly Rep. 2019;68(11):267–272. doi:10.15585/mmwr.mm6811e1

13. Eisinger RW, Dieffenbach CW, Fauci AS. HIV viral load and transmissibility of HIV infection undetectable equals untransmittable. JAMA - J Am Med Assoc. 2019;321(5):451–452. doi:10.1001/jama.2018.21167

14. El-Nahal WG, Shen NM, Keruly JC, et al. Telemedicine and visit completion among people with HIV during the coronavirus disease 2019 pandemic compared with prepandemic. AIDS. 2022;36(3):355–362. doi:10.1097/QAD.0000000000003119

15. Wood BR, Lan KF, Tao Y, et al. Visit Trends and Factors Associated With Telemedicine Uptake Among Persons With HIV During the COVID-19 Pandemic. Open forum Infect Dis. 2021;8(11). doi:10.1093/OFID/OFAB480

16. Boshara AI, Patton ME, Hunt BR, Glick N, Johnson AK. Supporting Retention in HIV Care: Comparing In-Person and Telehealth Visits in a Chicago-Based Infectious Disease Clinic. AIDS Behav. Published online February 3, 2022:1-7. doi:10.1007/S10461-022-03604-W/TABLES/2

17. Harsono D, Deng Y, Chung S, et al. Experiences with Telemedicine for HIV Care During the COVID-19 Pandemic: A Mixed-Methods Study. AIDS Behav. 2022;26(6):2099–2111. doi:10.1007/S10461-021-03556-7/TABLES/4

18. Galaviz KI, Shah NS, Gutierrez M, et al. Patient Experiences with Telemedicine for HIV Care During the First COVID-19 Wave in Atlanta, Georgia. https://home.liebertpub.com/aid. xPublished online May 11, 2022. doi:10.1089/AID.2021.0109

19. Grove M, Brown LL, Knudsen HK, Martin EG, Garner BR. Employing telehealth within HIV care: advantages, challenges, and recommendations. AIDS. 2021;35(8):1328–1330. doi:10.1097/QAD.0000000000002892

20. Wood BR, Young JD, Abdel-Massih RC, et al. Advancing Digital Health Equity: A Policy Paper of the Infectious Diseases Society of America and the HIV Medicine Association. Clin Infect Dis An Off Publ Infect Dis Soc Am. 2021;72(6):913–919. doi:10.1093/CID/CIAA1525

21. Budak JZ, Scott JD, Dhanireddy S, Wood BR. The Impact of COVID-19 on HIV Care Provided via Telemedicine—Past, Present, and Future. Curr HIV/AIDS Rep. 2021;18(2):98. doi:10.1007/S11904-021-00543-4

22. Mgbako O, Miller EH, Santoro AF, et al. COVID-19, Telemedicine, and Patient Empowerment in HIV Care and Research. AIDS Behav. 2020;24(7):1990–1993. doi:10.1007/s10461-020-02926-x

23. Smith E, Badowski ME. <p>Telemedicine for HIV Care: Current Status and Future Prospects</p>. HIV/AIDS - Res Palliat Care. 2021;13:651–656. doi:10.2147/HIV.S277893

24. Moore RD. Understanding the clinical and economic outcomes of HIV therapy: The Johns Hopkins HIV clinical practice cohort. J Acquir Immune Defic Syndr Hum Retrovirology. 1998;17(SUPPL. 1):S38–41. doi:10.1097/00042560-199801001-00011

25. A Timeline of COVID-19 Vaccine Developments in 2021. Accessed May 31, 2022. https://www.ajmc.com/view/a-timeline-of-covid-19-vaccine-developments-in-2021

26. CDX Technologies. Zip Code Distance Batch Report. Accessed May 31, 2022. https://www.cdxtech.com/tools/bulk/distance/

27. Kind AJH, Buckingham WR. Making Neighborhood-Disadvantage Metrics Accessible — The Neighborhood Atlas. N Engl J Med. 2018;378(26):2456–2458. doi:10.1056/NEJMP1802313/SUPPL_FILE/NEJMP1802313_DISCLOSURES.PDF

28. University of Wisconsin School of Medicine and Public Health. 2019 Area Deprivation Index. Accessed May 31, 2022. https://www.neighborhoodatlas.medicine.wisc.edu/

29. Olatosi B, Weissman S, Zhang J, Chen S, Haider MR, Li X. Neighborhood Matters: Impact on time living with detectable viral load for new adult HIV diagnoses in South Carolina. AIDS Behav. 2020;24(4):1266. doi:10.1007/S10461-019-02734-Y

30. Edmonds A, Breskin A, Cole SR, et al. Poverty, Deprivation, and Mortality Risk Among Women With HIV in the United States. Epidemiology. 2021;32(6):877–885. doi:10.1097/EDE.0000000000001409

31. Gu A, Yoo H Il. vcemway: A one-stop solution for robust inference with multiway clustering: https://doi.org/101177/1536867X19893637. 2019;19(4):900–912. doi:10.1177/1536867X19893637

32. Wood BR, Young JD, Abdel-Massih RC, et al. Advancing Digital Health Equity: A Policy Paper of the Infectious Diseases Society of America and the HIV Medicine Association. Clin Infect Dis. 2021;72(6):913–919. doi:10.1093/cid/ciaa1525

33. D D, BN D, B L, M T, TP G. Exploring the Attitude of Patients with HIV About Using Telehealth for HIV Care. AIDS Patient Care STDS. 2020;34(4):166–172. doi:10.1089/APC.2019.0261

34. Roberts ET, Mehrotra A. Assessment of Disparities in Digital Access among Medicare Beneficiaries and Implications for Telemedicine. JAMA Intern Med. 2020;180(10):1386–1389. doi:10.1001/jamainternmed.2020.2666

35. Eyrich NW, Andino JJ, Fessell DP. Bridging the Digital Divide to Avoid Leaving the Most Vulnerable Behind. JAMA Surg. 2021;156(8):703–704. doi:10.1001/JAMASURG.2021.1143

36. Estacio EV, Whittle R, Protheroe J. The digital divide: Examining socio-demographic factors associated with health literacy, access and use of internet to seek health information. J Health Psychol. 2019;24(12):1668–1675. doi:10.1177/1359105317695429

37. Busch AB, Sugarman DE, Horvitz LE, Greenfield SF. Telemedicine for treating mental health and substance use disorders: reflections since the pandemic. Neuropsychopharmacol 2021 466. 2021;46(6):1068–1070. doi:10.1038/s41386-021-00960-4

38. Yang J, Landrum MB, Zhou L, Busch AB. Disparities in outpatient visits for mental health and/or substance use disorders during the COVID surge and partial reopening in Massachusetts. Gen Hosp Psychiatry. 2020;67:100–106. doi:10.1016/J.GENHOSPPSYCH.2020.09.004

39. Eruchalu CN, Bergmark RW, Smink DS, et al. Demographic Disparity in Use of Telemedicine for Ambulatory General Surgical Consultation During the COVID-19 Pandemic: Analysis of the Initial Public Health Emergency and Second Phase Periods. J Am Coll Surg. 2022;234(2):191–202. doi:10.1097/XCS.0000000000000030

40. Reed ME, Huang J, Graetz I, et al. Patient Characteristics Associated With Choosing a Telemedicine Visit vs Office Visit With the Same Primary Care Clinicians. JAMA Netw Open. 2020;3(6):e205873–e205873. doi:10.1001/JAMANETWORKOPEN.2020.5873

41. Eberly LA, Kallan MJ, Julien HM, et al. Patient Characteristics Associated With Telemedicine Access for Primary and Specialty Ambulatory Care During the COVID-19 Pandemic. JAMA Netw Open. 2020;3(12):e2031640–e2031640. doi:10.1001/JAMANETWORKOPEN.2020.31640

42. Caregiving in America | The National Alliance for Caregiving. Accessed May 7, 2021. https://www.caregiving.org/research/caregivingusa/

43. Addressing Gender Disparities in Transportation - Gender Policy Report. Accessed May 7, 2021. https://genderpolicyreport.umn.edu/addressing-gender-disparities-in-transportation/

44. COVID-19 Vaccine Monitor Dashboard | Kaiser Family Foundation. Kaiser Family Foundation. Accessed May 26, 2022. https://www.kff.org/coronavirus-covid-19/dashboard/kff-covid-19-vaccine-monitor-dashboard/

45. Dasigi J V, Gupta N, Hadi C. 895. Impact of Telemedicine on HIV Care and Prevention Services at an Academic Ryan White-Funded Clinic. Open Forum Infect Dis. 2021;8(Supplement_1):S539–S539. doi:10.1093/OFID/OFAB466.1090

46. Grubesic TH, Matisziw TC. On the use of ZIP codes and ZIP code tabulation areas (ZCTAs) for the spatial analysis of epidemiological data. Int J Health Geogr. 2006;5:58. doi:10.1186/1476-072X-5-58

